# Comparison of Intramuscular Pharmacological Treatment Options for Acute Agitation: A Multicenter Retrospective Cohort Study

**DOI:** 10.1101/2025.06.17.25329649

**Authors:** Ananya Sri Thiriveedhi, Joshua Chang

## Abstract

**Objective:** Acute agitation in psychiatric settings presents significant clinical and safety challenges. Pharmacological management is often necessary when de-escalation strategies fail, but optimal medication regimens remain unclear. This study evaluates the safety, efficacy, and adverse event profiles of commonly used intramuscular (IM) pharmacologic regimens for acute agitation in a multicenter cohort.

**Methods:** We conducted a retrospective cohort study using de-identified data extracted from the HCA Healthcare corporate database. Adult psychiatric patients who received PRN intramuscular medications for acute agitation were included. Patients were stratified into four treatment groups based on administered medication: (1) haloperidol monotherapy, (2) haloperidol + lorazepam, (3) haloperidol + diphenhydramine, and (4) haloperidol + lorazepam + diphenhydramine. Outcomes assessed included frequency of PRN use, benztropine administration, and incidence of hypotensive and hypoxic episodes. Generalized linear modeling was used for statistical analysis.

**Results:** The combination regimen of haloperidol + lorazepam + diphenhydramine (Group 4) was associated with significantly higher odds of receiving multiple PRN administrations compared to haloperidol alone (Group 1). However, this regimen, along with the diphenhydramine-inclusive group (Group 3), was linked to a significantly lower likelihood of requiring benztropine, suggesting a reduction in extrapyramidal symptom burden. No statistically significant group differences were observed in hypotensive or hypoxic episodes.

**Conclusion:** Triple-agent regimens may be associated with increased treatment intensity but offer benefits in reducing extrapyramidal symptoms without increasing cardiovascular or respiratory risk. These findings support the thoughtful use of combination IM regimens, particularly those including diphenhydramine, in the management of acute agitation in psychiatric settings.

## Introduction

Acute agitation is a psychiatric emergency frequently encountered in inpatient behavioral health and emergency department settings. It is commonly associated with underlying psychiatric conditions, including schizophrenia, bipolar I disorder, and schizoaffective disorder, and may be exacerbated by environmental triggers, medication noncompliance, substance use, or psychosocial stressors. When not promptly and effectively managed, agitation can lead to harm to self or others, staff injury, and need for physical restraint or seclusion—all of which are associated with long-term psychological trauma and increased healthcare costs. Evidence-based management strategies recommend verbal de-escalation and environmental interventions as first-line approaches. However, pharmacologic treatment becomes essential when non-pharmacologic interventions are ineffective or unsafe. Despite its frequent use, there is no universal consensus on an optimal pharmacologic strategy. Haloperidol, a first-generation antipsychotic, remains a common first-line agent due to its rapid onset of action (typically within 15–60 minutes) and established efficacy. However, its potent dopamine D2 antagonism increases the risk of extrapyramidal symptoms (EPS), such as acute dystonia and akathisia, often requiring prophylactic or reactive treatment with anticholinergic agents such as benztropine. To mitigate these side effects, combination regimens including benzodiazepines or diphenhydramine are frequently used. Benzodiazepines, such as lorazepam, offer anxiolytic and sedative properties, while diphenhydramine, an antihistamine with anticholinergic properties, may help prevent EPS. Nevertheless, these combinations may also increase sedation or cardiorespiratory complications, and data comparing the safety and efficacy of these regimens remain sparse. A 2022 retrospective cohort study by Jeffers et al. offered some comparative insights but did not evaluate adverse events such as hypotension or hypoxia. This study seeks to address this knowledge gap by analyzing real-world data from multiple behavioral health facilities to assess how different IM medication regimens affect treatment outcomes in acute agitation, including frequency of medication administration, side effect mitigation, and cardiorespiratory safety.

## Methods

### Study Design and Population

This was a secondary analysis of a multicenter, retrospective cohort using de-identified data from the HCA Healthcare corporate database. Included were adult patients (≥18 years) admitted to behavioral health facilities who received PRN intramuscular medication for acute agitation between 2020 and 2023. Patients were required to have received at least one IM medication administration to be included in the sample.

### Treatment Groups

Patients were categorized into one of four groups based on the IM medication regimen administered during an agitation episode:

1. **Group 1:** Haloperidol only
2. **Group 2:** Haloperidol + Lorazepam
3. **Group 3:** Haloperidol + Diphenhydramine
4. **Group 4:** Haloperidol + Lorazepam + Diphenhydramine

### Outcomes Measured

Primary and secondary outcomes were compared between groups:

1. **Frequency of PRN medication administration:** Proxy for treatment efficacy or need for redosing.
2. **Benztropine administration:** Used as a marker for extrapyramidal symptoms.
3. **Hypotensive episodes:** Systolic blood pressure <90 mmHg.
4. **Hypoxic episodes:** Oxygen saturation <90%.

### Statistical Analysis

Outcomes were analyzed using generalized linear models:

- **Zero-inflated Poisson regression** was used to account for overdispersion and the excess of zeros in PRN count data.
- **Logistic regression with Firth’s penalized likelihood** was applied to reduce small-sample bias in binary outcomes such as benztropine use and adverse events.

## Results

### PRN Medication Frequency

Group 4 (haloperidol + lorazepam + diphenhydramine) had significantly higher odds of requiring more than one PRN administration compared to Group 1 (OR = 1.394, p < 0.001). No significant differences were observed between Groups 2 or 3 and Group 1, suggesting the increase in PRN usage was specific to the triple-agent regimen.

### Benztropine Administration

Group 3 (haloperidol + diphenhydramine) showed a statistically significant reduction in the likelihood of benztropine administration (OR = 0.549, p = 0.001), as did Group 4 (OR = 0.290, p< 0.001). This supports the hypothesis that diphenhydramine reduces EPS burden, potentially through its anticholinergic properties.

### Hypotensive and Hypoxic Episodes

No statistically significant group-wise differences were observed in hypotensive or hypoxic episodes. This finding suggests that combining lorazepam or diphenhydramine with haloperidol does not elevate the risk of cardiopulmonary adverse events within the defined post-administration window.

## Discussion

This study offers several clinically relevant insights into the pharmacologic management of acute agitation. While the triple drug combination (Group 4) was associated with increased PRN usage, possibly reflecting either lower efficacy in controlling agitation or a higher baseline severity among the patients, it was also associated with the lowest need for benztropine, suggesting a reduced risk of extrapyramidal symptoms (EPS). This trade-off may be acceptable in patients where EPS prevention is a priority, especially those with known EPS history or high D2-sensitivity. The observed safety profile was consistent across all medication groups. Neither the inclusion of lorazepam nor diphenhydramine increased the risk of hypotension or hypoxia, challenging concerns about over-sedation or respiratory depression commonly associated with these agents in combination. While no statistically significant group-wise differences were observed in cardiovascular and respiratory outcomes, it is possible that patients in the triple-drug combination group (Group 4) were more medically stable at baseline. This may reflect a selection bias, as geriatric patients with greater cardiovascular or respiratory instability were likely excluded from receiving the triple-drug regimen. Furthermore, this study has limitations inherent to retrospective designs. Confounding variables such as severity of agitation, prior medication history, or comorbid substance use could not be fully controlled. Furthermore, the right-skewed distribution of PRN counts posed analytical challenges, though advanced modeling techniques were used to mitigate this. The requirement that all patients must have received at least one administration also may have introduced selection bias, particularly excluding patients successfully managed without pharmacologic intervention.Future research should build upon these findings with prospective designs. Randomized controlled trials comparing monotherapy to dual- and triple-drug regimens, while adjusting for severity of agitation and psychiatric diagnosis, could validate and expand upon these findings. Additionally, studies comparing medication performance in the emergency department versus inpatient behavioral health settings may illuminate setting-specific practices.

### Conclusion

In this multicenter retrospective cohort study, diphenhydramine-inclusive regimens demonstrated a favorable side effect profile with reduced need for benztropine, without elevating cardiovascular or respiratory risk. While triple-agent regimens may be associated with increased PRN usage, they appear safe and potentially more tolerable in terms of EPS prevention. These data support the inclusion of diphenhydramine in IM treatment regimens for acute agitation, particularly for patients at high risk of EPS.

## Data Availability

All data produced in the present study are available upon reasonable request to the authors

## Notes

### Competing Interest Statement

The authors have declared no competing interest.

### Funding Statement

This study did not receive any funding

### Author Declarations

IRB of HCA Healthcare gave ethical approval for this work

